# Using Facebook groups to support families: midwives’ perceptions and experiences of professional social media use

**DOI:** 10.1101/2022.04.12.22273813

**Authors:** Holly Morse, Amy Brown

## Abstract

Seeking support from Facebook groups during pregnancy is now widespread and social media has been widely used by maternity services during the COVID-19 pandemic. Despite this, little is currently known about midwives’ attitudes towards, and experiences of social media in practice. Research is needed to understand barriers and solutions to meeting mothers’ expectations of online support and to improve services.

This study explored midwife involvement in Facebook groups, exploring experiences and perceptions of its use to communicate with and support mothers. 719 midwives and student midwives completed an online survey during August-September 2020 and their numerical and free-text responses analysed descriptively.

Few participants were involved in providing Facebook support, and most of these were unpaid. There was a consensus on a range of benefits for mothers, but widespread concern that engaging with mothers online was a personal and professional risk, underpinned by a lack of support. Experience of being involved in midwife moderation increased belief in its benefits and reduced fear of engaging online, despite a lack of renumeration and resources. Midwives and students felt they were discouraged from offering Facebook support and sought further training, guidance and support.

Although limited, experiences of providing Facebook group support are positive. Perceptions of risk and a lack of support are significant barriers to midwives’ involvement in using Facebook groups to support mothers. Midwives seek support and training to safely and effectively engage with mothers using Facebook.

Engaging with mothers via social media is embedded in national policy and digital strategy, and progress is needed to fulfil these, to improve services and meet mothers’ expectations. Midwives’ experiences suggest extending opportunities to provide Facebook support would benefit midwives, services and families. Consultation to revise local policy to support midwives and students in line with strategic goals is recommended.

**Author Summary:** Social media use continues to grow and we know that use among pregnant and new parents for peer support and accessing information is widespread. Previous research suggests however that they can find it difficult to know which information to trust, and would like to engage with their midwives online. However, little was known about how many midwives are supporting families via social media, or what their experiences of this are. Nor did we know what the perceptions of developing this service are amongst the workforce. Here, we used an online survey to explore these attitudes and perceptions. We found that although few midwives are engaging with families on social media, those that do have positive experiences. Many fear that a lack of support and guidance presents risks to themselves and to families, but recognised the potential benefits to developing the service. We also found that midwives would like to receive more training to deliver services online and engage safely on social media. Our study provides new insights that can be used to improve support for midwives and to realise the potential of social media in midwifery care.

## Introduction

There is a high level of public demand for responsive, evidence-based online health services [1] and the potential for these to improve care and increase efficiency is well documented [2]. Despite strategic emphasis on the digital transformation of the NHS, progress remains slow, particularly within maternity services [3]. However, digital technology was quickly utilised by services to play a vital role in disseminating information and communicating with families in response to the outbreak of COVID-19 [4], and work is needed to evaluate and maintain advances [3].

The popularity of Facebook support groups with pregnant and new mothers has grown rapidly in line with the increasing social media use amongst this demographic [5]. This use of peer-led online communities offers access to shared experience and social and emotional support that women find invaluable during the transition to parenthood [6]. However, whilst highly valuing them for social support, they encounter issues with whether the health information shared within online groups has any credible evidence base [7]. Inaccuracy and the sharing of misinformation within online groups is common which can heighten anxiety and increase help seeking from professionals [8,9]. Professionally mediated support offers a solution: facilitating peer support, relationship building and knowledge acquisition within an online community whilst addressing any inaccurate or misleading information [10]. Mothers seek this online support from midwives, and it improves their experience of midwifery care, and feedback on local maternity services [11].

Engagement on social media by NHS health professionals, to inform and empower service users, has been deemed critical for over a decade [12,2]. To facilitate this, sectors like pharmacy and dentistry have seen a growth in research into the issues surrounding digital professionalism [13,14]. The skills and professionalism required by nurses engaging online has more recently been explored [15,16], and training introduced to pre-registration programmes [17]. However, there is reluctance amongst midwifery educators to adopt such programmes [18]. Limited research suggests perceptions of the risks amongst midwives, and a lack of training present significant barriers to developing social media support more widely in maternity services [19,16].

The provision of midwife moderated Facebook groups offers maternity services an opportunity to develop support and health engagement, improving outcomes and experiences [20,11]. However, little is known about how midwives perceive and experience this form of communication and support, and no literature has been identified that explores any training or support offered to midwives related to this developing role. This study therefore aimed to explore midwives’ and student midwives’ attitudes towards, and any experiences of, delivering support to mothers via Facebook. Developing an understanding of barriers to developing this provision will help inform practice and education, supporting midwives to engage safely and effectively with digital support services. Specifically, the study sought to explore:

1. How are midwives using Facebook and what are their perceptions of professional social media use?
2. What are midwives’ experiences of, and concerns about offering social media support?
3. What training needs relating to social media and breastfeeding support do midwives have?

## Results

### Participants

Seven hundred and nineteen midwives and student midwives completed the online questionnaire. The participants fell into all provided age ranges, from 18 years to over 60 years (mean age range 22-30, median 31-40 years). The majority of participants identified as either White or White British (93%) and female (98.9%). This reflects the demographics of UK midwives [21]. Further details can be found in Table 1. Participants were asked to provide details of how long they had been qualified as a midwife (or were currently a student) and to indicate their current role. At the time of survey completion, over a third (36.2%, n = 260) were student midwives. For qualified midwives (n = 459), time since registration ranged from 0-20+ years, with 63.1% having been qualified 10 years or less. Overall, twice as many qualified midwives were based in hospital (30.2%) as were based in a community setting (clinic/home based care) (14.7%). Those with specialist roles (16.4%) were asked to give details, with infant feeding being the most commonly specified role (13.6%). A chi square found that those in specialist roles were significantly more likely to be providing Facebook support than those who were not in specialist roles [X^2^ = 20.067, p = .000].

**Table 1.** Demographics of respondents.

| Indicator | Group | N | % |
| --- | --- | --- | --- |
| Age | 18-21 | 73 | 10.4 |
|  | 22-30 | 229 | 32.6 |
|  | 31-40 | 207 | 29.4 |
|  | 41-50 | 118 | 16.8 |
|  | 51-60 | 64 | 8.9 |
|  | 60+ | 12 | 1.7 |
| Gender | Female | 695 | 98.9 |
|  | Male | 6 | 0.9 |
|  | Self-defined | 2 | 0.3 |
| Ethnicity | Asian or Asian British [Indian, Bangladeshi, Pakistani, Other] | 5 | 0.6 |
|  | Chinese | 0 | 0 |
|  | Black/Black British | 10 | 1.4 |
|  | Irish | 11 | 1.6 |
|  | Mixed or multiple | 18 | 2.6 |
|  | White/White British | 654 | 93.0 |
|  | Other | 5 | 0.7 |

### Facebook use

Participants were asked to indicate whether they used Facebook and type of use [none, personal use and/or connecting with other professionals, to offer women support], selecting all that applied. Personal use and connecting with other professionals was the most common type of use (41%, n = 295), followed by personal use only (36.4%, n = 262). Overall, 103 (14.3%) used Facebook to professionally support pregnant/postnatal women, usually alongside other personal and professional use (74.8%, n= 77) and 1.3% (n = 9) did not use Facebook. Participants who reported using Facebook were split into two groups for further analysis: those providing Facebook support (14.6%, n= 100) and personal/social use combined (85.4%, n = 584).

Using a five-point Likert scale [strongly agree to strongly disagree], participants were asked to indicate how they felt about a series of statements, including their trust in Facebook, whether significant people in their life were Facebook users, whether Facebook helped them learn from other professionals and if Facebook support could improve care for women. These results were compared for the two groups [personal/social and support]. Providing support was significantly associated with perceptions of trust, connection and improvements in care, confidence in online professionalism and being happy interacting with mothers online (Table 2).

**Table 2.** Statements on Facebook use.

| Type of Facebook use | Personal/Social |  | Support |  |  |
| --- | --- | --- | --- | --- | --- |
|  | Strongly Agree/Agree |  | Strongly Agree/Agree |  | Significance |
| Perception of FB use | N | % | N | % |  |
| I trust FB with my information | 146 | 25.9 | 37 | 41.6 | t [650] = 3.596, p = .000 |
| Enables me to connect/learn from other professionals | 449 | 80.2 | 83 | 93.3 | t [647] = 3.948, p = .000 |
| Friends/family are FB users | 429 | 76.3 | 67 | 75.3 | t [649] = .742, p = .459 |
| Social benefits | 482 | 85.8 | 78 | 87.6 | t [649] = 1.134, p = .257 |
| Convenient & easy to use | 540 | 96.1 | 83 | 93.3 | t [649] = .231, p = .818 |
| Confident in staying professional online | 480 | 85.4 | 84 | 94.4 | t [649] = 2.793, p = .005 |
| Happy to interact with mothers | 178 | 31.7 | 75 | 84.3 | t [649] = 10.311, p = .000 |
| Facebook use can improve care | 268 | 47.7 | 81 | 91.0 | t [649] = 8.818, p = .000 |
| Also use other social media | 440 | 78.3 | 68 | 76.4 | t [649] = 649, p = .073 |

### Facebook support roles

All participants were asked if their NHS health board/trust had official Facebook groups used by midwives to support local women. Of 517 responses, 296 (57.3%) did, 140 (27.1%) did not and 81 (15.7%) were unsure. Of those providing online support 63 (72.4%) were aware of or involved in their local NHS affiliated group. Participants were also asked if they had only started providing a Facebook support role as a result of the COVID-19 pandemic. This was the case for 13 (14.8%) participants. Those participants not providing support were asked if they would consider the role in future. Overall, of 393 completed responses, 56% felt they would or may, and 44% indicated they would not.

Participants with involvement in Facebook support indicated what their group offered, and their responsibilities in relation to the role. A combination of antenatal and postnatal support, including breastfeeding support, was most common (36.4 %, n = 32), 14 (15.9%) provided specialist support e.g. for NICU or parents of multiples and 13 (14.8%) were breastfeeding support groups. In relation to responsibilities, contributing to discussion by posting and responding to women’s posts alone was the most common (27.3%) followed by involvement in setting up the group, moderating discussion and responding to posts (22.7%). Overall, 18.2% (n = 16) were involved in discussion and moderation alone and 20.5% (n = 18) specified other responsibilities using a free text box. These included responsibility for promotion of their NHS or independent services using Facebook pages (rather than support groups), running support groups for professionals/students and involvement in digital intervention projects.

Participants were asked to indicate whether they were required to offer Facebook support as part of their employed role as a midwife. Overall 31.0% (n = 27) did so within their role, and (62.1%, n = 54) chose to do so outside of their employed role. The remainder (n = 7) were student midwives. Participants were also asked how many hours they spent on their Facebook role in an average week and what proportion, if any, they were paid for. Two to four hours was the most common weekly time spent on the role (35.1%, n = 26) with 4.1% (n = 3) spending over 30 hours. The majority were offering support outside of their employed role and not being paid for it (73.6%, n= 39) and only 32% (n=8) of those employed to offer Facebook support were fully reimbursed.

When asked how long they had been involved in providing Facebook support, most participants had been doing this role between 1-3 years (37.5%, n = 33), with 12.5% (n = 11) having started within the previous three months (during the pandemic), and 27.5% (n= 22) for over three years. Of those offering support as part of their employed role, most had been doing so under one year (59.2%, n = 16). Those midwives offering Facebook support outside their role had most often been doing so over a year (77.4%, n = 41).

### Perceptions of Facebook support

Participants were asked to rate a series of statements of positive impacts of mothers’ and midwives’ use of Facebook support groups using a five-point Likert scale [strongly agree to strongly disagree]. These statements focused on elements of knowledge acquisition and social capital. These responses were compared for the two types of Facebook use (personal/social and support (Table 3). On mothers’ use, there was a consensus of agreement with all statements, and strong agreement with the ability of Facebook support to provide peer support (95.3%) and access to shared experience (97.4%). Participants were less likely to agree with positive impacts on continuity of care (50.2%) and improvements in breastfeeding rates (60.6%). Participants who provided Facebook support reported significantly greater agreement with all statements, including improved feedback, communication, self-efficacy, confidence and knowledge (p = <0.05).

**Table 3.**
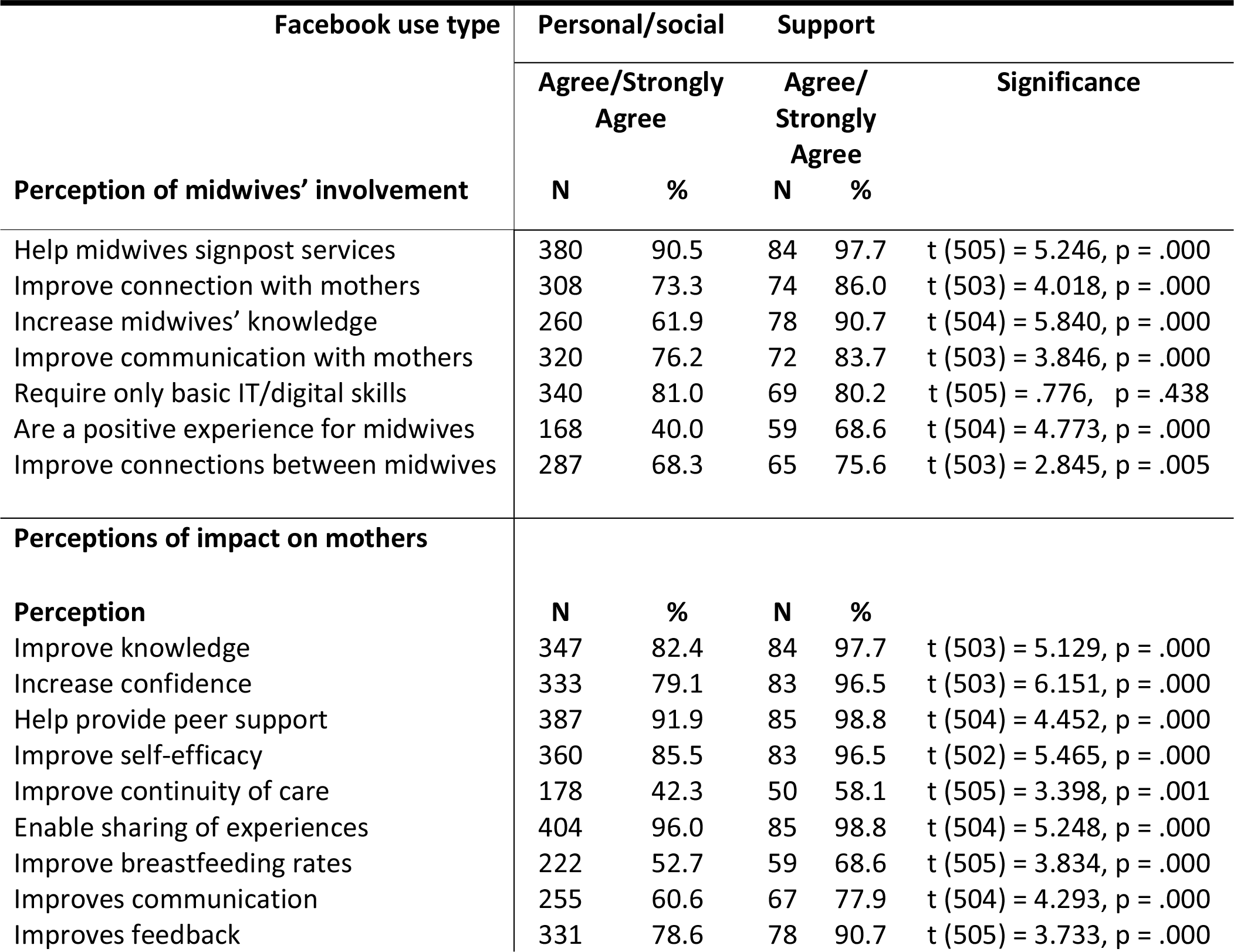
Perceptions of Facebook support group use.

Participants were also asked to rate a series of statements about midwives’ professional use of Facebook support groups, using a five-point Likert scale [strongly agree to strongly disagree]. For only basic IT skills being needed, those in the support group were less likely to strongly agree (M = 2.02, SD =.958), although this was not significant (p = .438). Participants providing support reported significantly greater agreement with all other statements (p = <.0.05).

### Concerns about providing Facebook support

All participants were asked to indicate whether and to what extent a list of personal and professional issues were of concern to them in relation to providing Facebook support as a midwife [very concerned - not a concern] (Table 4). The personal/social group reported significantly greater concerns compared to support providers for personal privacy, increased workload/stress, becoming emotionally involved and overstepping boundaries.

**Table 4:** Concerns about providing FB group support.

| Facebook use type |  | Personal/social |  | Support |  | Significance |
| --- | --- | --- | --- | --- | --- | --- |
|  |  | Concerned<br>N | % | Concerned<br>N | % |  |
| Personal | Personal privacy | 338 | 81.1 | 52 | 61.2 | t [505] = -4.090, p = .000 |
|  | Digital competence | 147 | 35.3 | 31 | 36.5 | t [504] = .449, p = .653 |
|  | Criticism from colleagues | 260 | 62.4 | 38 | 44.7 | t [505] = -2.625, p = .009 |
|  | Ensuring my advice is evidence based | 301 | 72.2 | 60 | 70.6 | t [503] = -.999, p = .318 |
|  | Increased workload/stress | 274 | 65.7 | 48 | 56.5 | t [502] = -3.266, p = .001 |
|  | Becoming emotionally involved | 254 | 60.9 | 39 | 45.9 | t [501] = -3.597, p = .000 |
|  | Overstepping boundaries | 324 | 77.7 | 42 | 49.4 | t [501] = -6.387, p = .000 |
| Professional | Posting publicly in case of error/complaint | 355 | 85.1 | 47 | 55.3 | t [503] = -5.662, p = .000 |
|  | Being reported to the NMC/my employer | 305 | 73.1 | 34 | 40.0 | t [500] = -6.222, p = .000 |
|  | Ensuring mothers' confidentiality | 311 | 74.6 | 43 | 50.6 | t [500] = -4.719, p = .000 |
|  | Lack of guidance for moderating groups | 351 | 84.2 | 44 | 51.8 | t [500] = -7.529, p = .000 |
|  | Lack of employer support | 317 | 76.0 | 44 | 51.8 | t [499] = -4.782, p = .000 |
|  | Managing conflict online | 347 | 83.2 | 44 | 51.8 | t [501] = -7.165, p = .000 |

There was a consensus of opinion on all areas of professional concern, with lack of guidance for moderating groups and public posting for fear of error or complaint being most strongly felt. Other concerns included being reported to the NMC/employer, lack of employer support and managing online conflict. Overall around two thirds to three quarters of the personal/social group held professional concerns compared to around half of those in the support group.

### Training and support for Facebook roles

Participants were asked to indicate whether they had received any training relating to social media use, and if they would find this useful. Few participants had received any relevant training. Any training was perceived as potentially useful by the majority of participants, whether they had received any training in the past or not. Of those who were providing Facebook group support, 9 (10.8%) had received digital skills training, 15 (18.1%) had received social media training and 15 (18.1%) had received e-professionalism training. Being involved in Facebook support was not significantly associated with having received any training. Just 22.2% of those providing support had received written local guidelines for their role, with 76.1% believing these would be useful.

Participants were asked whether they felt any of a list of professional and managerial sources were supportive of midwives providing Facebook support to mothers (Table 5). Overall, work colleagues were seen as supportive most often and NHS health board/trust management the least. Those involved in providing support reported greater perceptions of support for the role, than the personal/social group.

**Table 5:** Perceptions of professional support for FB group roles.

| Facebook use type | Personal/<br>social |  | Support |  |  |
| --- | --- | --- | --- | --- | --- |
|  | Agree/ Strongly<br>Agree |  | Agree/Strongly<br>Agree |  | Significance |
| Source | N | % | N | % |  |
| Facebook group guidelines | 133 | 32.2 | 44 | 51.8 | t [498] = 3.298, p = .001 |
| Health board/NHS trust<br>Management | 114 | 27.6 | 30 | 35.3 | t [494] = 2.617, p = .009 |
| NMC social media guidance | 167 | 40.4 | 44 | 51.8 | t [494] = 1.941, p = .053 |
| The Royal College of Midwives | 181 | 43.8 | 40 | 47.1 | t [495] = 1.053, p = .293 |
| Work Colleagues | 176 | 42.6 | 53 | 62.4 | t [494] = 2.180, p = .030 |
| Universities/<br>midwifery educators | 175 | 42.4 | 37 | 43.5 | t [492] = -.579, p = .093 |

Participants provided further detail on their views, concerns, reasoning and experiences via a free text box. Thematic analysis identified three overarching themes: Progress, Threat and Barriers.

#### 1. Progress

The concept of Facebook support as progress, encompassed perceptions of positive, or potential positive impacts on the maternity service, on women and on midwives. Some participants described the provision of midwife led Facebook support as improving maternity services and care for women. This included increased engagement, improved feedback and the development of services to meet strategic and service user expectations, particularly in the context of the COVID-19 pandemic.

> *“A positive has been that women have shared their birth stories with thanks and appreciation for the great care they have received-this improved morale for the midwives and provided other mums to be with some reassurance.”* (Birth centre midwife)
>
> *“We are in an age of social media. People expect to be able to use these methods to communicate. As service providers we need to be able to adapt and use the same platforms as our service users to ensure we provide evidence based and accessible care.”* (Community midwife)

The use of Facebook groups to offer both professional and peer support within an online community, particularly where in person support had been affected by the coronavirus pandemic was also seen as positive. This encompassed antenatal, postnatal and infant feeding and included informational and social support.

> *“The sense of community on social media platforms between expectant mothers has been a lot more apparent during lockdown as women have been unable to attend face to face parentcraft/mother and baby classes etc and have been reaching out to each other a lot more online for advice and just general friendship.”* (Student midwife)

Midwives had observed positive impacts on self-efficacy and improved relational continuity.

> *“Women have felt a little put out during this pandemic and I have noticed a lot of activity on a local health board website for feeding. It has been great to watch others peer support each other and midwives to continue with continuity that otherwise could have been lost.”* (Hospital midwife)

Participants described a belief in the potential of Facebook support to improve care and experiences for mothers and midwives, including breaking down barriers and continuity of care.

> *“I’d love to (give FB support)! Think it would bring down a lot of the unequal power dynamics that are implicit by being a HCP.”* (Hospital midwife)

Several described how it improved their own job satisfaction by enabling relationship building.

> *“Women who have had support from me on my personal midwife Facebook page, allow me to provide continuity of care and achieve a sense of fulfilment from being a midwife that my job no longer gives.”* (Community midwife)

#### 2. Threat

The concept of Facebook support as a being a ‘threat’ encompassed perceptions of potential negative impacts or risks to the maternity service, on women and on midwives. Participants described how engaging with Facebook created opportunities for negative feedback and challenges in managing responses, which impacted personal wellbeing and wider staff morale.

> *“It’s a great idea however, trust is then up for public verbal abuse and becomes frustrating when you can’t challenge the ‘keyboard warriors’.”* (Hospital midwife)

Offering support via Facebook was also considered a potential threat to women’s access to individualised information and care, or as a way to justify reductions in service provision. Participants feared being held accountable for this.

> *“*[Social media support was*] seen as temporary during extremis. When all face-to-face meetings can resume the concern is that they will not - women will be left with inadequate levels of care and midwives held responsible for mopping up issues via Facebook. It’s another NHS care scandal waiting to happen.”* (Hospital midwife)

Participants also described concerns that social media groups pose a threat to appropriate and effective information sharing, perpetuating false information with the potential for adverse outcomes.

> *“It’s impossible to gather all the information [in a Facebook group], other non-medical professionals give their opinion diluting the impact of the health care professionals’ advice.”* (Student midwife)

Some perceived the use of Facebook as a threat to women’s relationships with services/care providers, and the ability of professionals to communicate effectively and individually with them. There were beliefs that Facebook support groups offered false security and an obstacle to appropriate help-seeking.

> *“Unfortunately [social media use] does mean there is a delay in some women accessing appropriate care, as they will message with a serious concern when the inbox is not manned (though it is widely publicised what times it is manned!)”* (Community midwife)

Participants had concerns about upholding their personal boundaries and the potential for the time, privacy and wellbeing of midwives to be threatened by engaging on social media.

> *“It’s difficult when you are seeing bad advice being given by others. I have also seen some quite unprofessional responses from the moderators of one NHS run group which only served to fuel the fire…Then it feels almost personal when someone attacks as they are talking about you and your work colleagues.”* (Hospital midwife)

Experiences highlighting issues of professionalism, appropriate engagement and defensive practice, threatening professional reputations and wellbeing were shared. Often the solution to these issues was seen as withdrawing from any public engagement.

> *“Any advice given by midwives can and will be screenshotted, edited, shared on women’s personal accounts, and has huge potential personal risk to midwives. I have seen many midwives berated and destroyed on social media, comments taken out of context… I don’t think any interactions should be public”*. (Hospital midwife)

#### 3. Barriers

Three distinct ‘Barrier’ themes to the use of social media to provide support were identified: fear, lack of training and lack of support. Fears were centred on the implications of engaging publicly on their status as a registered midwife, lack of guidance and clarity on appropriate use and accountability.

> *“I would be mindful that my philosophy of practice does not marry with the expectations of some employers. I’d also be concerned about being judged by others who do not share similar approaches to care. I would feel I’d have to “tow the line” in that I could only offer support and advice that is aligned with institutional midwifery care.”* (Midwife)

Fears relating to security and confidentiality, as well as the potential for being judged personally and professionally in public were widely held. Student midwives felt a lack of support through their education and were impacted by the conflicting attitudes.

> *“Our lecturers have given us extremes of views on social media use… it has been confusing. Most of my cohort are like me, very worried about what we post online to the point where we probably wouldn’t. This would mean women miss out on that advice/connection. I think we need more training other than scaremongering. I think the NMC should have really clear-cut advice about…whether students/midwives should be using social media to connect and when it’s appropriate.”* (Student midwife)

Participants described how a lack of training designed to support and clarify the use of social media to enhance care presented a barrier to safe and confident use, cementing fears around professionalism.

> *“I think we need to move away from the assumption social media=\=unprofessional, and all the training I’ve had or discussed with others have been about protecting yourself and defensiveness, not about safe usage or recommendations to improve care.”* (Community midwife)

It was also widely felt that there is a lack of support, hostility and resistance from employers, professional bodies and universities, presenting a barrier to Facebook to provide support.

> *“I’ve only ever sought to connect women with services when they cannot reach them, correct misinformation, and reach out when there is clear distress. My employer responded by threatening me with referral to NMC for using Facebook on trust time and misrepresenting the trust online.”* (Hospital midwife)

## Discussion

This study explored midwives’ and student midwives’ (referred to in the discussion jointly as ‘midwives’) attitudes toward, and experiences, of using Facebook to provide midwifery support. A growing body of research shows that women expect health services to use digital tools for support and communication, and that social media has the potential to improve these services [22,2]. Our study explored the barriers to this provision within maternity services, from the perspective of midwives. Consistent with other research [19,16], the findings show midwives are concerned about the risks of offering support to pregnant and new mothers online and seek more training and guidance to do so. However, those delivering support via Facebook viewed the role and its impact positively and had fewer concerns, despite a lack of guidance and resources. These findings have important implications for developing the guidance and support to safely deliver progress in utilising social media as a professional tool.

Seeking support on Facebook during pregnancy and early parenting is now widespread and has a wide range of benefits for mothers. Facebook support groups offer convenient access to highly valued social and emotional peer support, increasing confidence, knowledge acquisition and self-efficacy [23]. Midwives recognised that many mothers are now using Facebook in this way but were also aware that mothers have issues with recognising reliable information, and that the sharing of unhelpful, and, in some cases risky, advice is a common experience [8]. Our findings showed that many midwives saw midwife-moderation as a solution to validating information in Facebook groups, which is reflected in the literature [10]. However, others felt that this created or may create opportunities to undermine individual midwife-mother relationships and for mothers to challenge those providing their clinical care. This was not the experience of those actually involved in Facebook support, who viewed interactions as positive for mothers, midwives and services. These findings are important for considering how to incorporate midwives’ experiences into training that addresses concerns and supports service development.

Belonging to a Facebook support group provides mothers with a support network and access to lived experience that is not always available to them within their local community. Groups that offer support to mothers located within specific geographic areas enable ‘real life’ connections to be built, fostering relationships that create social opportunities for mothers and babies and facilitating signposting to face to face support when needed [11].\ These mothers also have shared experiences of their maternity services and expect to receive updates and offer feedback on their care via social media. Midwives had mixed experiences of this, sharing the boost to morale of appreciation and positive engagement, but also the impact of negative interactions, particularly in the context of changes related to COVID-19 [24]. Faced with inaccuracies, judgement or criticism about maternity care or services on Facebook midwives felt disempowered, and that they were did not have the guidance or support to engage constructively. These negative experiences on Facebook in general impacted midwives’ attitudes towards Facebook support, despite not being seen in midwife-moderated groups [10]. Previous research found access to midwife moderated groups improved perceptions of midwifery support, improving feedback and experiences, including during the pandemic [25]. Findings suggest training should be developed to support professional and constructive interactions which promote positive relationships between families and services.

There was a consensus amongst midwives that relevant skills and knowledge training was lacking and would be useful, particularly in relation to digital professionalism. Midwives are socialised through their education and practice to understand what is expected of them [26], and the standards they must uphold to register, and remain registered, with the NMC [27]. This extends to online behaviours and interactions [28], and although midwives felt they were confident in staying professional online, they were less so about maintaining boundaries and how to ensure confidentiality. This underpinned midwives’ fears that engaging online could prompt complaints and referral to their employer or the NMC, despite this being highly infrequent [18]. Universities were also seen as unsupportive; student midwives reported being warned against social media use during their education and felt frustration at defensive rather than proactive approaches. Previous research has noted resistance to introducing digital professionalism to the midwifery curriculum [18]. These findings are a concern, highlighting that midwives are often not receiving the support or training needed, pre- or post-registration, to meet digital transformation goals.

Although health professionals have long been encouraged to engage with service users on social media [12] midwives felt prevented from doing so by local policies and a lack of employer support. This is in line with previous research, which identified that whilst NHS strategy calls for greater engagement, professionals are discouraged by a focus on security and reputation [29]. Many midwives perceived any interaction with mothers on Facebook as inappropriate and unprofessional and most felt NMC [30] social media guidance did not support midwives to provide Facebook support. This reflects the generic referencing within the guidance of relationship building as unprofessional and inappropriate, despite improved relational continuity being a key benefit of midwife moderated Facebook groups [20]. These findings are important to consider in relation to the development of online support services, updating training and policies to reflect national strategy and the evidence base supporting this provision.

Midwives involved in providing support experienced improved connections with mothers and providing continuity as personally fulfilling, as well as recognising the benefit of continuity to mothers’ wellbeing and pregnancy outcomes [31]. Midwives felt their knowledge had increased as a result of being involved in moderating an online community, learning from mothers’ questions and experiences. Health professional moderators commonly report increased learning and research opportunities, and that the role can be personally and professionally empowering [32]. Midwives in the personal use only group were less likely to perceive these benefits, or the potential for engaging online to support continuing professional development. Findings suggest this may be a missed opportunity and that widening access to midwife moderator roles would benefit individuals and services.

Midwives expressed fears for their own privacy, security and a desire to avoid blurring boundaries between personal and professional online space. Social media use requires energy and cognitive processing that can cause overload and fatigue [33], and additional professional use may increase these risks, creating anxiety, stress and rumination [34,35]. There was concern about differentiating between being ‘on’ and ‘off’ duty when an online workplace is accessible around the clock and carried in a pocket. They feared becoming emotionally involved and overstepping boundaries. Some shared experiences of midwives being identified or targeted on Facebook, or having information they had shared taken out of context, creating fear this would occur in a group setting. However, these concerns were significantly associated with those not already involved in offering support, indicating that group formats and guidelines can support mutually respectful interactions [20]. Notably, around a third of those offering support online had concerns about doing so, suggesting their belief in the benefits motivated them to manage their concerns/accept a level of perceived risk in order to offer this support. Further research is needed to understand how personal and professional social media ‘roles’ and ‘profiles’ can be delineated to protect midwives and their wellbeing and promote effective practice.

Participants were also concerned about the potential increased workload and stress arising from a social media role. Linked to this are wider issues of the systemic undervaluing of midwives work and skills [36], including regularly missed breaks and unpaid overtime [37]. Existing understaffing has been compounded by Brexit and the pandemic, with Heads of Midwifery reporting that services frequently rely on the goodwill of staff to keep going [36]. Fair pay has been an ongoing issue over the last decade, with the value of midwives’ wages decreasing in real terms by over £7000 since 2010 [38]. The systemic undervaluing of their work and skills has exacerbated low morale, contributing to almost three quarters of midwives considering permanently leaving the profession [36]. Few midwives providing Facebook support were being paid to do so, even where this was part of their employed role. The majority were doing so outside an employed role and in their own time, and most had no local guidelines to support the role. One participant described the sudden acknowledgement of social media support during the COVID-19 pandemic as worthy of paid time, as ‘a kick in the guts’, expressing the frustration of those midwives seeking recognition for the time and skills they invest in this [and all] provision. Social media roles, where there are no guidelines, working hours or renumeration in place, clearly carry a risk of exacerbating existing work-based inequities and increasing pressures.

These findings are a concern, demonstrating a failure to support, safeguard and renumerate midwives who provide a service that mothers seek and services benefit from [11]. In addition to the potential personal and professional risks being shouldered by midwives, this situation also prevents effective auditing of any midwife-led online support to ensure its quality, efficacy, safety and accessibility. Being able to identify who is moderating a group is key to how mothers engage with and perceive its reliability [10,11]. It is therefore vital that services ensure midwife moderated groups are part of a robust, professional and accountable digital service [2].

While midwives in the sample often presented binary views of Facebook support as a benefit or a threat to care and services, they were aware that women’s expectations in relation to digital communication are changing. However, concerns were expressed that social media support would be relied upon to replace and justifying reduced resourcing of face-to-face services. Clearly new approaches to offering support will be needed, but these must be based on evidence, meet mothers’ needs and be integrated with care into the role of the midwife. The findings support wider research, highlighting a desire to understand ‘cybercivility’ [appropriate online engagement] and develop skills in digital professionalism [39]. This need has been brought into focus by the rapid digitalisation of services during COVID-19 [24]. Overall, findings highlight that further training is needed, and that midwives are eager to engage where guidance exists, and where policy supports practice.

The research does have limitations. This was an exploratory study in a new area which relied on large scale recruitment online, attracting participation amongst midwives and student midwives via social media posts and online sharing. Whilst efficient, this recruitment method meant those midwives active on social media may be more likely to respond and will have attracted those most motivated to take part. Although efforts were made to share the link to participate via the RCM channels and sharing encouraged outside social media were encouraged, those who choose not to use social media are less likely to have been represented. Just 1.3% of study participants were non-Facebook users, compared to 33% of the UK population, and 14.5% of 25-40 year olds [40]. Whilst this may be partially a result of the internet data collection methods, the demographic of midwives and student midwives in the sample reflected the childbearing women they care for [the majority being female and aged 22-40 years], who themselves represent the largest number of social media users [5]. It is unsurprising therefore that almost all participants were active Facebook users, that 95.1% find the platform convenient and easy to use and that 77.6% also used other social media such as Twitter or Instagram. Limitations of data collection methods aside, these findings suggest that much of the midwifery workforce is familiar with using social media for personal use, potentially providing a strong foundation for developing skills for the midwifery moderator role. However, many participants also expressed strongly held views about the personal nature of their Facebook use and a desire to keep professional life and engagement with women separate. Overall, 44% stated they would not consider a role that involved offering Facebook support in future. It is evident that digital skills present less of a barrier than the pervasive conception of social media use as unprofessional.

Similarly, although our sample was predominantly from White or White British backgrounds [93%], this number and the representation of other ethnic groups reflects the number of registered midwives from these backgrounds in the UK [21].

The questionnaire design relied on self-reports and although anonymous, social desirability bias should be considered in survey responses by professionals [41]. Social media use by midwives is presented as professionally problematic by educators, employers and professional bodies, which could lead to denying involvement or exaggerating negative views. However, participants were self-selecting and questions carefully worded to minimise any bias.

Limitations aside, this study has demonstrated that there are significant perceived personal and professional barriers to the integration of Facebook as a tool for supporting mothers into midwifery practice. Despite strategic goals encouraging social media interaction and the impact of the COVID-19 pandemic on delivering digital communication, progress in maternity services remains slow. Mixed messages between local and national policies and defensive social media policies are causing fear amongst midwives. A lack of support from employers and resistance from midwifery educators is preventing creative approaches to overcoming the complexities of using Facebook to support families. It is clear midwife moderated Facebook support has the potential to support skill development, improve communication and meet women’s needs. However, the application of the knowledge, skill and passion of midwives to delivering support via social media needs wider exploration to ensure access is equitable, appropriately resourced and midwives are supported, protected and renumerated. Further research also needs to establish how appropriate digital professionalism training can be developed and implemented to reduce fear and improve engagement. This is vital if maternity services are to meet mothers’ expectations for digital access to support, and strategic goals for digital transformation.

## Materials & methods

### Participants

The sample was a convenience sample recruited online between 1^st^ August and 30^th^ September 2020. Inclusion criteria were: aged over 18 or over, a registered midwife or student midwife in the UK and who gave consent to participate in the survey in English. Responses were received from regions across the UK. Ethical approval was granted by a University Research Ethics Committee. All participants gave informed consent to take part in the study.

### Questionnaire Design

An exploratory online survey, consisting of open and closed questions, was used to collect data on the attitudes of midwives towards Facebook use, the benefits and challenges of developing the midwife’s role in Facebook group provision, and barriers to development of the service. Participants completed an online questionnaire asking them about their experiences and/or perceptions of the use of Facebook to provide mothers with support. The questionnaire included items exploring:

- Age, gender and ethnicity. Participants also gave employment details including whether they were currently a student, or how long they had been a midwife and any specialist roles. County area was also collected to determine the geographic spread of participants.
- Measures of Facebook use: including type of use and perceptions of use.
- Format of Facebook support roles: including types of support, responsibilities held, time spent and reimbursement.
- Perceptions of Facebook support roles: including any additional breastfeeding qualifications, perceptions of impact of groups on mothers and midwives.
- Training and support for Facebook roles: including any training and/or guidelines received or perceived as needed.

The questionnaire was piloted prior to sharing more widely. It was completed by 3 midwives and five student midwives. Feedback from initial participants was positive on structure and content. No changes were required.

### Procedure

Participants were recruited to the study using an advertisement with a link to the online questionnaire, hosted by Qualtrics. Facebook groups aimed at midwives and students [such as ‘Beyond Midwifery UK’ and ‘Midwives in the making’] were identified via a Facebook search, with permission sought from group administrators for posting study information to the group or page. The advertisement and link were shared to these groups and to midwifery related Facebook pages and shared by members and on the Royal College of Midwives [RCM] website. It was also shared more broadly across social media. If participants were interested in taking part, they clicked on the link where the participant information sheet and consent questions loaded. A short debrief was included at the end of the questionnaire with details of how to contact the research team or seek further support if needed.

### Data Analysis

Data were descriptively analysed using frequencies and percentages using SPSS v26. Participants were asked to indicate whether they used Facebook [Not at all, personal/social use, to provide professional support to women]. Non-users were directed only to questions on views, omitting use and experiences. As the research questions focus on understanding the experiences of Facebook users [and the non-user sample very small], non-user responses were excluded from some analyses. Where non-users are included, results refer to all participants.

Chi square tests were carried out to compute associations between type of Facebook use [personal/social use, professional support use] and age range, specialist role, group recommendations and receipt of training. T tests were performed to compare attitudes to mothers and midwives’ use of Facebook support groups and level of concern about their use for the two Facebook use groups. County area data were analysed for distribution frequency using Google My Maps.

Thematic analysis was conducted to explore patterns and connections within the qualitative data. After familiarisation with the data, initial codes were produced, identifying themes which were reviewed in relation to the coded extracts, defined and named. These were reviewed by a second researcher and discussed until agreement reached [42]. A reflexive journal was used to reflect on methodological decisions and the researcher’s midwifery background. Results were audited by the second researcher, providing feedback on the adequacy of data, development of findings and the interpretive perspective [43].

## Data Availability

All data underlying the findings reported are provided as part of the submitted article.

